# COVID-19: disease pathways and gene expression changes predict methylprednisolone can improve outcome in severe cases

**DOI:** 10.1101/2020.05.06.20076687

**Authors:** Sorin Draghici, Tuan-Minh Nguyen, Larry A. Sonna, Cordelia Ziraldo, Radu Vanciu, Raef Fadel, Austin Morrison, Mayur Ramesh, Gil Mor

## Abstract

**Background:** Current management efforts of COVID-19 include: early diagnosis, use of antivirals and immune modulation. After the initial viral phase of the illness, identification of the patients developing cytokine storm syndrome is critical. Treatment of this hyper-inflammation in these patients using existing, approved therapies with proven safety profiles could address the immediate need to reduce the rising mortality.

**Methods:** Using data from an A549 cell line, primary human bronchial epithelial (NBHE), as well as from COVID-19-infected lung, we compare the changes in the gene expression, pathways and mechanisms between SARS-CoV2, influenza A, and respiratory syncytial virus.

**Results:** We identified FDA-approved drugs that could be repurposed to help COVID-19 patients with severe symptoms related to hyper-inflammation. An important finding is that drugs in the same class will not achieve similar effects. For instance **methylprednisolone** and **prednisolone** were predicted to be effective in reverting many of the changes triggered by COVID-19, while other closely related steroids, such as **prednisone** or **dexamethasone**, were not. An independent clinical study evaluated 213 subjects, 81 (38%) and 132 (62%) in pre-and post-methylprednisolone groups, respectively. The composite end point was composed of escalation to intensive care units, need for mechanical ventilation, and death. The composite endpoint occurred at a significantly lower rate in post-methylprednisolone group compared to pre-methylprednisolone group (34.9% vs. 54.3%, p=0.005).

**Conclusion:** Clinical results confirmed the efficacy of the *in silico* prediction that indicated methylprednisolone could improve outcomes in severe COVID-19. These findings are important for any future pandemic regardless of the virus.

## 1 Introduction

Most current efforts related to COVID-19 span a number of areas as follows: i) antivirals, ii) vaccine development, iii) diagnostic tests, iv) patient-supporting interventions. Without reducing the significance and impact of any of the areas above, there is an important aspect that has not been elucidated: the identification and treatment of patients developing critical conditions and risk of mortality. Mehta *et al*. state in a very recent Lancet paper [12]: “Accumulating evidence suggests that a subgroup of patients with severe COVID-19 might have a cytokine storm syndrome.” This cytokine syndrome correlates with high mortality. We propose that identification and appropriate management of the patients developing cytokine storm syndrome is critical for successful outcomes. Treatment of hyper-inflammation in these patients using existing, approved therapies with proven safety profiles could address the immediate need to reduce the rising mortality.

Unlike other efforts related to COVID-19, the work presented here focuses on: i) **understanding immunological response by lung epithelial cells to COVID-19 infection** and ii) **identifying drugs that would mitigate or alleviate some of the devastating over-reactions of the host’s immune system** (e.g. cytokine storm) that lead to poor outcomes, including death.

A SARS-CoV2-specific vaccine or a SARS-CoV2-specific antiviral will reduce the impact of this particular virus in future seasons. However, better understanding the acute reaction of the immune systems and having more tools to mitigate and/or avoid a cytokine storm will be **important for any future pandemic regardless of the virus**.

## 2 Results

We used available transcriptomic data to compare A549 lung cell line infected with SARS-CoV-2 vs. mock infection (henceforth A549CoV2vsMock), A549 infected with seasonal influenza A virus vs mock infection (A549IAVvsMock), and A549 infected with human respiratory syncytial virus vs mock infection (A549RSVvsMock). We also compared the transcriptional response in both primary human bronchial epithelial (NHBE) cells between cells infected with SARS-CoV2 and mock infection (NHBE-CoV2vsMock). Finally, we compared the transcriptional response in COVID-19 lung tissues vs. healthy lung tissue (COVID19vsHealthy). These data were collected at Mount Sinai and are available in GEO as the GSE147507 data set [4].

### Disrupted genes and biological processes

Fig. S1 shows a comparison of the affected biological processes in COVID19vsHealthy, NHBECoV2vsMock, A549CoV2vsMock, A549IAVvsMock, and A549RSVvsMock. The biological processes (BPs) are shown ordered by their significance in the COVID19vsHealthy. In spite of the larger number of DEGs in the COVID-19-infected lung (815), there are only 7 significant biological processes involved, which may indicate a more coordinated, systemic response. In contrast, the changes in the NHBE cells are characterized by fewer DEGs (only 223) but span more uncoordinated biological processes. This is illustrated in Fig. S2 which shows the BPs ordered in the order of significance in the NHBECoV2vsMock. Fig. S3 shows the Venn diagram representing the differentially expressed genes (DEGs) in the two contrasts. A comparison of the genes DE in the five contrasts is shown in Fig. S4.

### Putative mechanisms of disease

We performed an analysis aiming to identify putative mechanisms of disease. As part of this analysis we identified four genes that were predicted to be activated upstream regulators based on the observed changes in their downstream genes. These were IRF9, STAT2, IFNG and IFNB1. These suggest two different potential mechanisms. The first appears to be triggered by STAT2 and IRF9, which have 16 common target genes that are also all significantly up-regulated (IFI6, IFIT1, IFIT2, IFIT3, IFITM1, IFITM3, OAS1, OAS3, OAS2, MX1, MX2, RSAD2, OASL, XAF1, IRF2, IRF7). This mechanism is also known to be involved in the response to influenza A (see influenza A pathway in Fig. S5).

The second putative mechanism involves the interferon beta and gamma, which are targeting 5 common downstream genes: CXCL10, IDO1, DOX58, STAT1, which are up-regulated, and HMOX2 which is down-regulated. Interferon regulatory factors (IRFs) are subdivided into the interferonic IRFs (IRF2-3-7 and 9), the stress responsive IRFs (IRF1 and 5), the hematopoietic IRFs (IRF4 and 8) and morphogenic IRF-6. IRF9 is a regulator of type I IFN signaling and is known to interact with STAT2 [9] and STAT1 to form the heterotrimeric transcription factor complex (ISGF3) that binds to interferon-stimulated response elements (ISREs) to induce the expression of Interferon stimulated genes (ISG). During viral infections, ISGs perform two key functions: 1) directly limit viral replication by shutting down protein synthesis and triggering apoptosis; 2) ISGs activate key components of the innate and adaptive immune system, including antigen presentation and production of cytokines. The genes triggered by the STAT2 and IRF9 pathway include genes responsible for limiting viral replication (IFI6, IFIT1, IFIT2, IFIT3, IFITM1, IFITM3, OAS1, OAS3, OAS2, MX1, MX2, RSAD2, OASL) and inducers of apoptosis (XAF1, IRF2, IRF7). CXCL10, IDO1, DOX58, and STAT1 are genes associated with immune recruitment and immune regulation

Interestingly, STAT2 and IRF9 together are also identified as activated upstream regulators due to 15 downstream targets even in the NHBECoV2vsMock (see Fig. S6). However, in the NHBE cells, the interferon activators were replaced by an interleukin-based mechanism centered around IL1B, IL6, IL17A, adiponectin (ADIPOQ) and tumor necrosis factor (TNF).

We also looked at genes that are known to modulate or inhibit the inflammatory response such as IL1RN IL10, and IL13. In the COVID19vsHealthy, IL1RN was up 6.2 fold (FDR-corrected *p =* 10^−6^), IL10 was up 1.5 fold (FDR-corrected *p* = 0.55), while the measurement for IL13 was not available. In the NHBECoV2vsMock, IL1RN was up only 0.331 fold (FRD-corrected *p* = 0.035), while measurements for IL10 an IL13 were not available. However, in this contrast, 14 out 15 De genes immediately downstream of IL10 were up-regulated which strongly supports the hypothesis that IL10 is inhibited (FDR-corrected *p* = 5.17^−9^).

### Impacted pathways

The significantly impacted pathways are shown in Fig. S7 ordered by their significance in COVID19vsHealthy. The p-values represent a combination of enrichment and perturbation p-values (see [8] for details) corrected with FDR. Fig. S8 shows the signaling pathways in all 5 experiments, ordered by their significance in NHBECoV2vsMock.

Fig. S9 shows the most impacted pathway, the *Cytokine-cytokine interactions*. Fig. S10 shows the *Chemokine signaling pathway*. On this pathway, the impact is due both to the large number of DE genes (26 out of 130), as well as to the clear signal propagation from the chemokines outside the cell (11 chemokines up-regulated), through the chemokine receptor and via the JAK and STAT mechanism. Fig. S12 shows another view of the mechanism involving the genes on this pathway and all their known interactions.

### Proposed drugs

Once we identified the main regulatory pathways potentially associated with hyper-inflammation we evaluated *in silico* FDA-approved drugs that could show activity on multiple components of inflammation and consequently could be used for the management of severe COVID-19 cases. We considered the number of DE genes that would be reverted by each drug, as well as calculated a Bonferroni-corrected p-value indicating the suitability of each drug for repurposing in COVID-19 based on two different approaches (see methods). We looked for drugs that have both small p-values as well as revert a larger number of DE genes. The top three drugs drug identified by our analysis are shown in Fig. 1. **Methylprednisolone** is the drug that was identified as the most likely to work. This drug targets 27 genes that are found to be DE in COVID19vsHealthy. Out of these 27 genes, the drug would revert the changes in 25 of them. The drug also had an extremely significant p-value (*p* = 5.72^-10^) even after a Bonferroni correction which is the most stringent correction available. Methylprednisolone also reverted 22 out of 22 genes found to be DE in NHBECoV2vsMock, and 25 out of 26 genes found to be DE in A549CoV2vsMock. Fig. 2 shows the mechanism through methylprednisolone acts on the DE genes in COVID19vsHealthy, and how these genes influence the BPs found to be significantly impacted.

**Figure 1:**
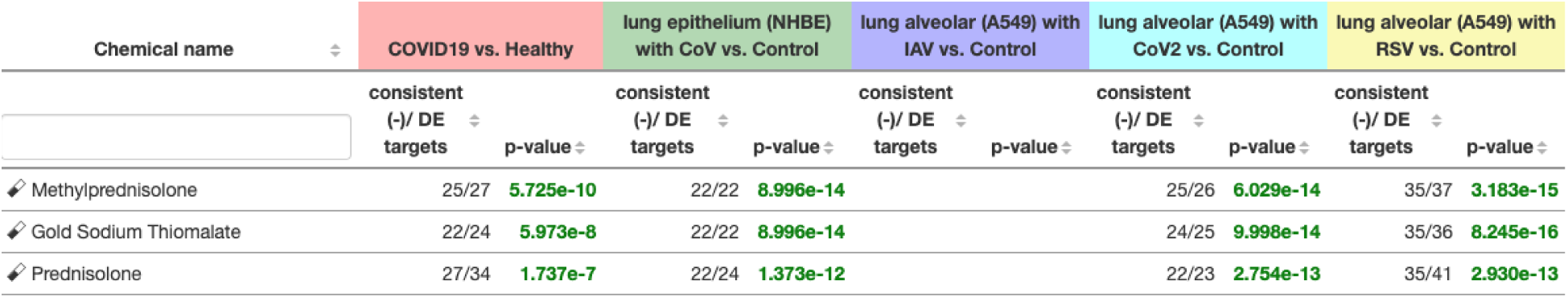
The top three drugs proposed for repurposing. The table shows both p-values corrected with Bonferroni, as well as the number of DE genes that would be reverted by each drug. Prednisolone and methylprednisolone are steroids currently used to modulate the immune response in rheumatoid arthritis. The column for A549IAVvsMock is empty because there are no DE genes targeted by these drugs in this contrast.

**Figure 2:**
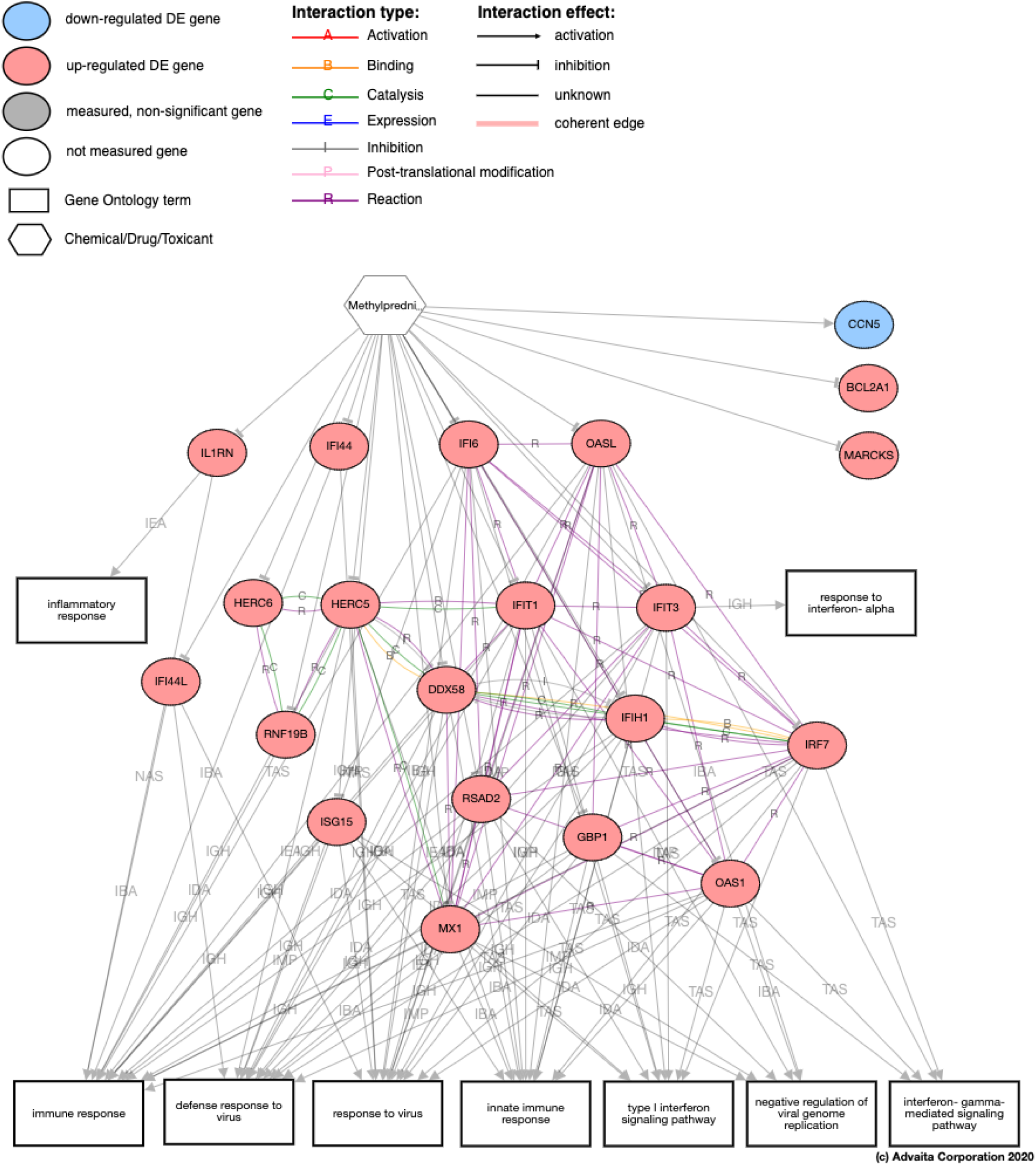
The mechanism through which methylprednisolone act on the genes measured to be DE and how these genes influence the biological processes found to be significantly impacted in the COVID19vsHealthy. Note how most of these genes are implicated in one or more of the dys-regulated biological processes.

### Clinical validation

An independent pre-treatment, post-treatment experiment was performed in a multi-center health center in Michigan between 12-27 March, 2020 in hospitalized patients with moderate to severe COVID-19. The protocol included an early, short-course of methylprednisolone: .5 to 1 mg/kg/day divided in 2 intravenous doses for 3 days. The study used a quasi-experimental design with 213 eligible subjects, 81 (38%) and 132 (62%) in pre-and post-corticosteroid groups, respectively. The primary end point was composed of escalation to ICU, need for mechanical ventilation, and death. The composite endpoint occurred at a significantly lower rate in post-methylprednisolone group compared to pre-methylprednisolone group (34.9% vs. 54.3%, p=0.005). The treatment effect was observed for each individual component of the composite endpoint. Significant reduction in median hospital length of stay was observed in the post-methylprednisolone group (8 vs. 5 days, p < 0.001). There was also an independent reduction in the composite endpoint at 14-days controlling for other factors (OR=0.45: 95% CI [0.25-0.81]). The survival curves of the pre-protocol and methylprednisolone groups are shown in Fig. 3. The clinical characteristics of the patients are shown in Table S2.

**Figure 3:**
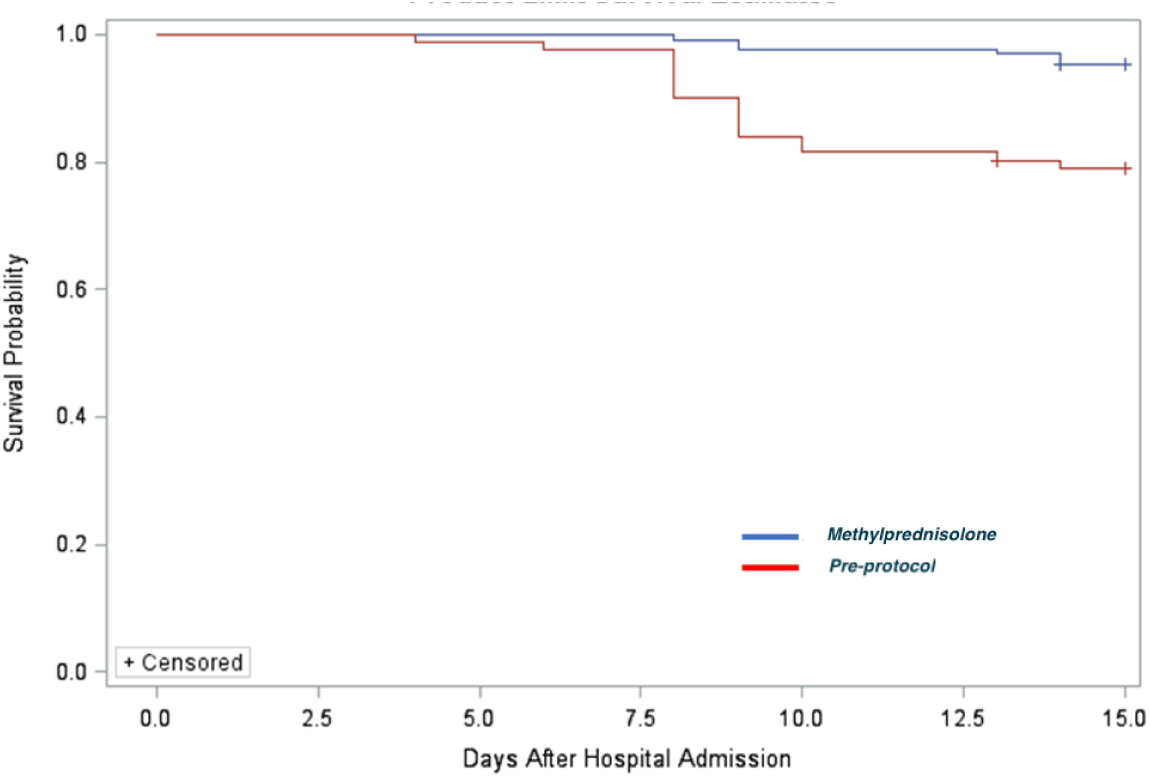
Survival curves comparing the evolution of patients treated with the standard of care (pre-protocol) and with methylprednisolone.

## 3 Discussion

Two recent papers stress the importance of a clinical phenotyping that would distinguish the phase where the viral pathogenicity is dominant versus when the host inflammatory response overtakes the pathology [3,14]. A strong argument in favor of also targeting the host response is offered by the data on influenza. Even though influenza patients receive optimal anti-viral therapy, approximately 25% of the critically ill influenza patients still die [3,11]. This suggests that anti-virals alone will not be sufficient for COVID-19 either, and the host response to the virus still needs to be taken into consideration.

However, approaches aiming at modulating the immune response face some concerns. In particular, it may seem counter-intuitive to try to diminish the immune response in a patient whose immune system is fighting against a virus. Modulating the immune system is likely unnecessary and counterproductive for patients whose immune system is doing a good job at resolving the infection, while it could potentially be life-saving for those whose inflammatory response has become dysregulated. If a patient has developed severe respiratory symptoms and is hypoxic, the host response that lead to ARDS, sepsis, and organ failure has already been initiated [12]. At this point, the focus should shift to supporting the patient’s systems and preventing collapse triggered by hyper-inflammation [3].

An important finding is that drugs in the same class will not have similar effects. For instance, **methylprednisolone** and **prednisolone** were predicted to be effective in reverting many of the changes triggered by COVID-19, while other closely-related steroids such as **prednisone** or **dexamethasone** were not. **Methylprednisolone** and **prednisolone** are steroids currently used to modulate the immune response in rheumatoid arthritis. The mechanisms through which these drugs would revert the genes dysregulated in COVID-19 are shown in 4. We also looked at other steroids such as **prednisone**, **dexamethasone**, and **hydrocortisone**. However, prednisone was found to target only 3 genes that are DE in the COVID19vsHealthy and only 2 of the genes that are DE in the NHBECoV2vsMock. From those, prednisone would revert only 1 of the 3 DE genes in the COVID19vsHealthy and 0 out of the 2 in the NHBECoV2vsMock. Both yielded insignificant p-values *(p* = 1 after Bonferroni) suggesting that prednisone is not expected to be an effective treatment. Prednisolone, dexamethasone, and hydrocortisone belong to the same family of corticosteroidal anti-inflammatory agents and there is also a structural similarity between them (see Fig. S14). In spite of this structural similarity, hydrocortisone would only revert 8 out of 10 genes found to be DE in the COVID19vsHealthy (*p* = 0.57 after FDR) and 5 out of 8 genes found to be DE in the NHBECoV2vsMock (*p* = 0.038 after FDR, *p* =1 after Bonferroni). Dexamethasone was found to revert 33 out of 69 of the genes found to be DE in the COVID19vsHealthy (*p* =1 after FDR correction) and 27 out of 45 genes in the NHBECoV2vsMock (*p* = 0.002 after fDr correction, *p* = 0.066 after Bonferroni correction). In short, neither dexamethasone nor hydrocortisone appears to be effective in the COVID-19 lung tissue, although hydrocortisone appears to be marginally effective in the NHBE.

**Figure 4:**
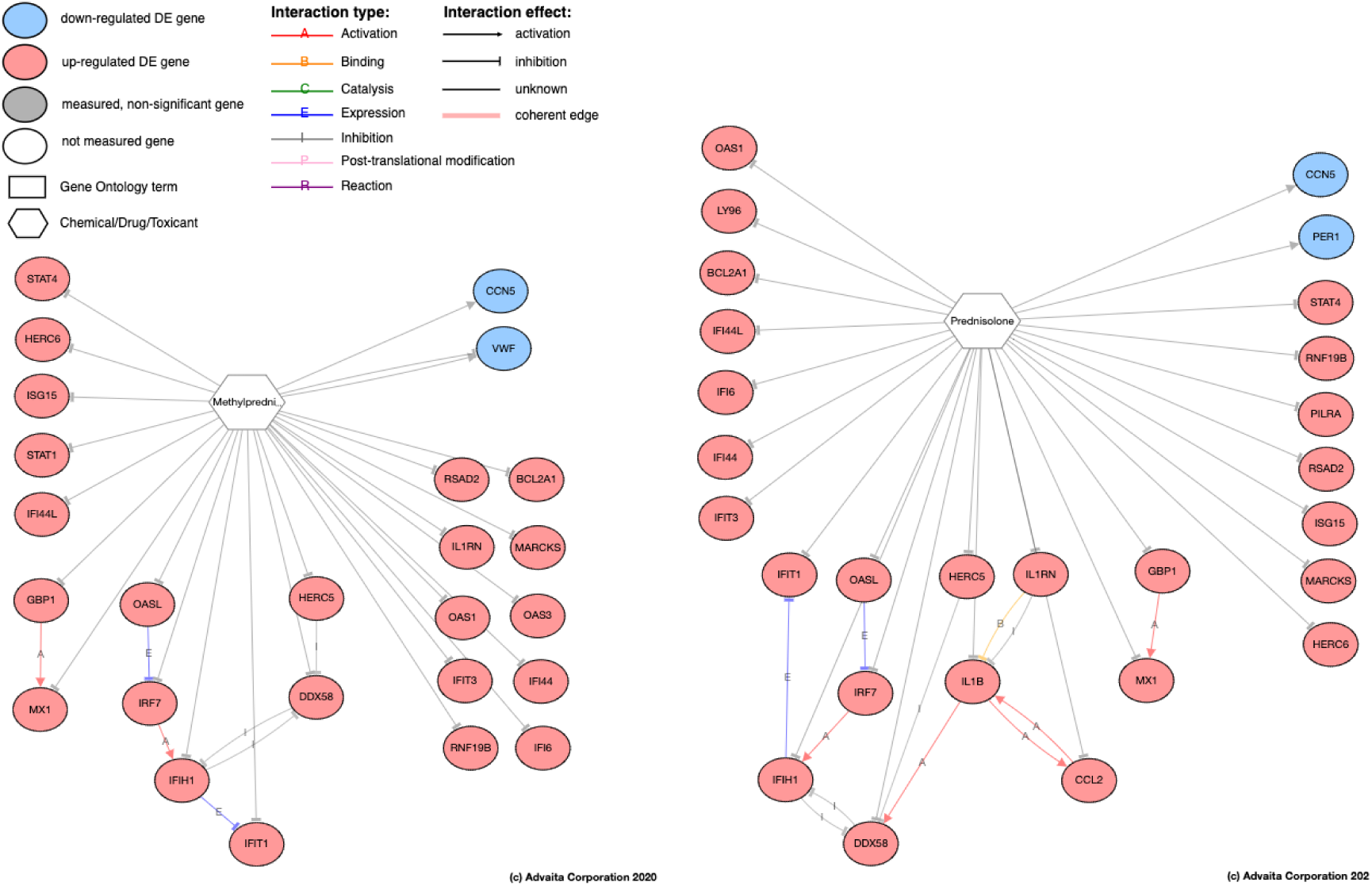
The putative mechanisms through which Methylprednisolone (left panel) and Prednisolone (right panel) would revert the changes triggered by COVID-19 in the lung tissue.

The host inflammatory response in the lungs may lead to acute lung injury and acute respiratory distress syndrome (ARDS). This constitutes the main rationale for potentially using corticosteroids. However, corticosteroids may have adverse effects, an increased risk of secondary infection and delayed viral clearance. A recent article in Lancet reports that clinical evidence does not support corticosteroid treatment for COVID-19 [13]. However, this report looks at steroids as an entire class of drugs. A recent retrospective study of 201 patients with COVID-19 in China found that treatment with methylprednisolone for those who developed ARDS was associated effective in decreasing the risk of death. Among patients with ARDS, treatment with methylprednisolone decreased the risk of death (HR, 0.38; 95% CI, 0.20-0.72). (23/50 [46%] with methylprednisolone vs 21/34 [17]. Both reports are entirely consistent with our findings: corticosteroids in general are NOT expected to help as a class of drugs. However, **methylprednisolone** and **prednisolone** are targeting a large number of the genes affected by COVID-19 and are expected to work significantly better than other corticosteroids.

We also looked at other drugs that have already been proposed as repurposing candidates for COVID-19 including: **chloroquine**, **hydroxychloroquine**, **erythromycin**, **prednisone**, **dexamethasone**, **ibuprofen**, **ritonavir**, **aspirin**, and **clopidogrel**. None of these was predicted to be effective (see Supplementary Materials).

## 4 Methods

The **primary endpoint** included: i) escalation to ICU, ii) progression to respiratory failure requiring mechanical ventilation, or iii) in-hospital all-cause mortality. Patients admitted directly in ICU were monitored for ii and iii above. Patients requiring mechanical ventilation upon admission were evaluated for mortality only.

### The methylprednisolone protocol

Moderate COVID-19 was treated with hydroxychloroquine 400 mg twice daily for 2 doses on day 1, followed by 200 mg twice daily on days 2-5. Patients with moderate COVID-19 who required 4 liters or more of oxygen per minute on admission, or who had escalating oxygen requirements from baseline, were recommended to receive IV methylprednisolone 0.5 to 1 mg/kg/day in 2 divided doses for 3 days. Patients who required ICU admission were recommended to receive the above regimen of hydroxychloroquine and IV methylprednisolone 0.5 to 1 mg/kg/day in 2 divided doses for 3 to 7 days. ICU patients were also evaluated for tocilizumab on a case-by-case basis. Oral switch was performed to prednisone at a ratio of 1 to 1 when determined clinically appropriate by the primary medical team.

### Statistical Analysis of clinical data

Continuous variables were reported as median and interquartile range (IQR) and compared using the Mann-Whitney test or t-test, as appropriate. Categorical data was reported as number and percentage (no., %) and compared using the chi-squared test or Fisher‘s exact test, as appropriate. No imputation was made for missing data points. The sample included all eligible consecutive hospitalized patients during the study period. More details about the statistical analysis and characteristics of the patient population are included in the Supplementary Materials.

### Pathway analysis method

iPathwayGuide (www.advaitabio.com) assesses pathways using the Impact Analysis method [8,10,15]. The impact analysis uses two types of evidence: i) the overrepresentation of differentially expressed (DE) genes in a given pathway and ii) the perturbation of that pathway computed by propagating the measured expression changes across the pathway topology. These aspects are captured by two independent probability values, *pORA* and *pAcc*, that are then combined in a unique pathway-specific p-value. More details are provided in the Supplementary Materials and elsewhere [8,16].

### Gene Ontology (GO) analysis method

For each GO term [2,5], the number of DE genes annotated to the term is compared to the number of DE genes expected just by chance. The p-value is computed using the hypergeometric distribution [6,7] and corrected with fDr and Bonferroni. We also used in intelligent prunning approach inspired by the *elim* and *weight* pruning methods [1]. The algorithm constructs a custom cut through the GO hierarchy by starting with the most specific nodes and calculating their p-value with all genes assigned directly to each such node. If a node is significant, it is reported as such. If the node is not significant, the genes associated to the given node are propagated to its direct ancestors and a p-value is calculated for each of those. See Supplementary Materials for full details.

### The prediction of upstream Chemicals, Drugs, Toxicants (CDTs)

is based on two types of information: i) the enrichment of DE genes from the experiment and ii) a network of interactions from the Advaita Knowledge Base (AKB v1910, www.advaitabio.com). The network is a directed graph in which the source node represents either a chemical substance or compound, a drug, or a toxicant (CDT). We focused our work on FDA-approved drugs that could be repurposed. The edges represent known increase or decrease expression effects that these CDTs have on various genes. The analysis considers the hypothesis that a drug would revert the measured gene expression changes. Full details are included in Supplementary Materials.

### 5 Conclusions

Clinical results confirmed the efficacy of the *in silico* prediction that indicated methylprednisolone could improve outcomes in severe COVID-19. These findings are important for any future pandemic regardless of the virus.

## Data Availability

The gene expression data analyzed here are available in GEO as data set GSE147507. All clinical data will be publicly available one year after the peer-review publication date.

